# A placental transcriptional signature for autism

**DOI:** 10.64898/2026.07.06.26357412

**Authors:** Luba Sominsky, Anne-Louise Ponsonby, Martin O’Hely, Richard Saffery, Christos Symeonides, Poshmaal Dhar, David Burgner, Peter D. Sly, Fiona Collier, Samuel Tanner, Katherine Drummond, Chloe J Love, Kristina Vacy, Toby Mansell, Sean L. McGee, Michael Berk, Peter Vuillermin, the Barwon Infant Study Investigator Group

## Abstract

Autism development involves multiple genetic and early-life environmental factors. Studying the placenta’s gene expression profile may reveal key mechanistic pathways in autism development. Here, using a nested case-cohort design within an Australian population-derived prebirth cohort study (n=1074), we identified 1,644 differentially expressed genes (DEGs; FDR<0.05) in the placenta of children with autism diagnosis (n=43), compared to those without (n=120). The top enriched pathways related to mitochondrial translation, oxidative stress, RNA processing and transcription regulation. *CYP1A1*, the most important xenobiotic-metabolising enzyme of the placenta, was the top downregulated DEG in the placenta of children with autism, while immuno-regulatory human leukocyte antigen (HLA)-related genes were among the top upregulated DEGs. A machine learning-based approach predicted autism from the transcriptomic data with a median sensitivity of 0.57 (2.5th–97.5th centiles: 0.29, 0.76) and median specificity of 0.92 (2.5th–97.5th centiles: 0.78, 0.98). Weighted Gene Correlation Network Analysis identified eight affected placental gene modules, with the largest five modules being enriched primarily for mitochondrial bioenergetics, oxidative phosphorylation and RNA processing pathways. This placental transcriptomic signature of impaired mitochondrial function and gene transcription regulation among infants subsequently diagnosed with autism has profound implications for understanding both risk factors and prediction, suggesting the possibility of identifying modifiable prenatal pathways to improve autism outcomes.

## Introduction

Autism is a complex neurodevelopmental condition characterised by impaired communication and repetitive behaviour^1^. It affects how a person interacts and communicates with others, as well as how they experience the world ^2^. Here we use the term *autism* rather than *autism spectrum disorder*, in keeping with preferences expressed by the autism community^3,4^. The number of individuals diagnosed with autism is increasing worldwide^5^. Between 2000 and 2020 the estimated prevalence of autism by 8 years of age in the United States increased four-fold from one in 150 to one in 36 children^1^, but the drivers of this increase remain unclear. Diagnostic changes seem to only partly explain this increase^6^. The aetiology of autism is complex, involving many genetic and environmental factors and their interplay^7^. Recent evidence indicates the placental transcriptome could be key to understanding genetic and early-life environmental factors in autism development^8,9^.

In mammalian pregnancy, the placenta is an indispensable transient organ, anchoring the embryo to the uterus and acting as a conduit between the environment, the mother and the developing foetus^10^. Placental pathology is associated with pregnancy complications and adverse foetal development, with the foetal brain being particularly vulnerable^11^. This vulnerability is due to the dynamic nature of placental adaptation to environmental challenges, affecting placental production of hormones, immune markers and neurotransmitters that influence foetal brain development. Increasing evidence indicates that disrupted placental function influences development of neurobehavioural disorders, including autism^11^. Differences in placental weight and morphology have been linked to autism-related traits and risk^12,13^. Placental insufficiency, vascular damage and enhanced systemic inflammation, often manifested in women with preeclampsia, are also associated with a higher risk of autism diagnosis^14^. These effects may result in part from limited availability of nutrients and oxygen for mitochondrial energy generation in the foetus, leading to progressive oxidative stress during critical periods of foetal brain development, as well as foetal growth restriction and progressive intrauterine hypoxia. It is notable in this context that the brain is the most energy dependent tissue in the body and that mitochondrial dysfunction primarily manifests with neurological symptoms^15^. The energy requirements are likely to be particularly high *in utero* when brain growth is rapid^16^. Knowledge of the role of placental biology in autism pathogenesis is likely to clarify operative pathophysiology and hence create opportunities for novel interventions to optimise prenatal brain development^17,18^.

Recent placental transcriptomic studies indicate that placental gene dysregulation partly mediates the association between prenatal environmental factors and adverse neurodevelopmental outcomes in children, including autism^8,9,19–22^. Placental DNA epigenetic studies have also identified methylation profiles that could be affected by maternal environmental exposures, to drive subsequent differential gene expression that leads to impairments in neurodevelopment and autism development^23,24^. These studies identified differential expression of several genes that are broadly associated with inflammation, immune response and neurodevelopment. However, the existing studies are relatively small, and no consistent predictive placental transcriptome signatures for autism have yet been identified.

The lack of placental function studies in relation to autism from diverse, large and well characterised cohorts remains an important research gap. To address this, we conducted a nested case-cohort study within a population-derived Australian pre-birth cohort (Barwon Infant Study (BIS); n=1074) comparing the placental transcriptional signatures and pathways among infants with a subsequent diagnosis of autism to a random sub-cohort of infants without autism diagnosis.

## Methods

### Participants characteristics

BIS is a population-based pre-birth cohort recruited using an unselected antenatal sampling frame in the Barwon region of Victoria, Australia (1064 mothers; 1074 children). The cohort details, inclusion and exclusion criteria have been previously described^25^. Information on preconception and antenatal periods was collected during the in-clinic appointment at 28 weeks gestation. The study was approved by the Barwon Health Human Research Ethics Committee (HREC 10/24), and families provided written informed consent.

Participant retention in the 9-year review was 85% (908/1074). Eighty parents reported their child had or was under assessment for autism, and two paediatricians from our team reviewed information abstracted from the medical records of 99% (79/80) of these children, to identify 64 children with a validated diagnosis of autism made by the child’s paediatrician against DSM-5 criteria by 11.5 years of age. Here, we used a case-cohort design to compare placental transcriptomes of children with autism for whom a validated diagnosis was available at the time of RNA-sequencing (RNA-seq) analysis (n = 43; 29 males/14 females), and a random sample of the cohort without autism (n = 120; 60 males/60 females). In the case-cohort analysis, as in our prior work^26^, children in the random sub-cohort with autism (n = 16) were classified as members of the case group. To provide a comparison to another developmental condition, we examined the placental transcriptome dataset of 52 children (29 males/23 females) with challenge-proven IgE-mediated food allergy at 1 year of age, compared to 115 (58 males/57 females) placental samples from a random sample of the BIS cohort.

### Placental tissue collection and RNA synthesis

Four full-thickness cores were taken from each placenta collected from 1013/1074 infants (1013/1064 mothers) shortly after birth, using an 8 mm biopsy punch^27^. Tissue samples were rinsed in phosphate buffered saline and stored in RNA*later* (Sigma-Aldrich, Castle Hill, Australia) at −80°C until use. To generate RNA, slivers from each placental biopsy were excised and homogenised separately in Buffer RLT (Qiagen, Hilden, Germany). For RNA synthesis, 100 μL from each homogenate were pooled and RNA was extracted using the QIAsymphony RNA Kit on the QIAsymphony SP instrument (Qiagen), according to the manufacturer’s instructions.

### mRNA Next Generation sequencing and analysis

Total RNA samples were submitted to the Australian Genome Research Facility (AGRF; Melbourne, VIC, Australia) for sequencing and bioinformatics analysis. RNA quality was assessed using an Agilent Bioanalyzer 2100. Samples were sequenced on an Illumina NovaSeq platform. The primary bioinformatics analysis involved demultiplexing and quality control (QC), with all samples passing QC. The data were then processed through an RNA-seq expression analysis workflow, which included trimming, alignment, transcript assembly, feature quantification and differential expression analysis, as previously described^28^.

The primary sequence data were generated using the Illumina’s Dynamic Read Analysis for GENomics (DRAGEN^tm^) Bio-IT Platform and DRAGEN BCL Convert 07.021.645.4.0.3 pipeline. The sequence reads from all samples were analysed according to AGRF quality control measures. The per-base sequence quality for the samples indicated excellent quality, with >89% bases above Q30 quality score across all samples. The reads were also screened for the presence of any Illumina adapter/overrepresented sequences and cross-species contamination.

The cleaned sequence reads were aligned against the *Homo sapiens* genome (Build version HG38). The STAR aligner (v2.3.5a) was used to map reads to the genomic sequences^29^. The counts of reads mapping to each known gene were summarised. The transcripts were assembled with the StringTie tool v2.1.4 (http://ccb.jhu.edu/software/stringtie/) utilising the reference annotation-based assembly option (RABT). This generated the assembly of both known and potentially novel transcripts. The RefSeq annotation containing both coding and non-coding annotations for human genome version hg38 was used as a guide (https://www.ncbi.nlm.nih.gov/assembly/GCF_000001405.39).

We used edgeR (Empirical Analysis of Digital Gene Expression Data in R, version 3.38.4) in R 4.2.2 to perform the differential expression analysis, as well as the gene ontology (GO), and Kyoto Encyclopedia of Genes and Genomes (KEGG) and Reactome pathway enrichment analysis. edgeR is a package used to detect and quantify differential expression of digital gene expression data, that is, counts of reads mapped for each gene of a given organism^30^. We used the default trimmed mean of M values (TMM) normalization method of edgeR to normalise the counts between samples^31^. A generalised linear model (using glmFit and glmLRT from edgeR) was then used to quantify the differential expression between the groups^32^. The differential expression analysis compared the autism case-group with the random sub-cohort. Foetal sex was included in the analysis model to reduce the impact of differences between male and female placenta, relating to metabolism, biosynthesis and immune function^33^. We also re-ran the differential expression analysis using only the placentas of male infants, and we ran the analysis with only foetal sex in the model to confirm that we were not simply seeing a signature of foetal sex. Gestational age was not included in the model as it may act causally downstream from the placental transcriptome and thus is not appropriate to control for.

### Ingenuity Pathway Analysis (IPA)

To interpret the DEGs in terms of the biological pathways and regulatory networks they participate in, we used Ingenuity Pathway Analysis (IPA; QIAGEN Inc., https://digitalinsights.qiagen.com/IPA) to perform the core analysis, exploring the canonical pathways, upstream regulators and causal networks among the DEGs using IPA default settings. To identify analysis-ready molecules, statistical significance was calculated using the right-tailed Fisher’s Exact Test. Benjamini-Hochberg correction for multiple comparisons was applied to calculate False Discovery Rate (FDR) < 0.05^34^. The canonical pathways functionality of IPA was used to determine which metabolic and cell-signalling pathways are predicted to be activated or inhibited based on significant enrichment of the DEGs. The activation/inhibition states of canonical pathways were predicted based on a z-score algorithm. Positive and negative z-scores represent activation and inhibition respectively; we applied a threshold z-score magnitude of at least 2, and *p* <0.05. The Upstream Regulator Analysis was used to identify the upstream molecules that are either activated or inhibited to directly drive the differential gene expression in the dataset, using the same thresholds (z-score magnitude ≥ 2; *p* < 0.05)^35^.

We then used enhanced Causal Network Analysis in IPA to identify master upstream regulators that control the expression of the DEGs in the dataset, either directly or through intermediate regulators. This analysis tool also allows assessment of connections between the master regulators and a particular downstream disease or biological process, and the role of target genes (DEGs) in these connections based on the Ingenuity Knowledge Base, a large collection of observations in various experimental contexts with nearly 5 million findings manually curated from the biomedical literature or integrated from third-party databases^35^. Here, we assessed the causal relationships with the following pre-defined conditions in IPA: ‘autism spectrum disorder or intellectual disability’ and ‘autism-like behavioural deficits.

### Prediction of autism DSM-5 diagnosis

In addition to differential expression and pathway analyses, we used our placental RNA-seq data to develop a machine-learning based prediction model of autism. We took counts-per-million values for the 16,478 genes which had been admitted to the DE analysis after edgeR’s default filtering and TMM normalisation, and included foetal sex as a predictor. We generated 100 instantiations of a 1:1 training:test split of the 43 cases and 120 children from the random sub-cohort from that analysis and built a sparse partial least squares discriminant analysis (sPLS-DA) model^36^ on each training set, using the implementation in the R package ‘spls’ (version 2.2-3; https://CRAN.R-project.org/package=spls) with default parameters and eta=0.7, K=10. We used this model to predict autism status in the corresponding test set, recording the confusion matrix for each instantiation and summarising via specificity and sensitivity. These were calculated using the confusion_matrix() function from the R package qwraps2^37^ and the Wilson score method for confidence intervals^38^. We repeated the above procedure using only predictor genes which were found to be differentially expressed in the edgeR analysis, acknowledging that this probably permits information leakage from the DE analysis.

### Weighted Gene Co-expression Network Analysis (WGCNA)

We then used WGCNA, an unsupervised, data-driven clustering technique, to identify gene modules based on shared expression. The input to WGCNA was the normalized expression values for the top 5000 DEGs (using TMM normalisation and p values from the edgeR differential abundance analysis above), selecting soft threshold power using pickSoftTreshold() from among whole numbers from 1 to 10 and even numbers from 12 to 20. The parameters used in blockwiseModules() were power as described above, maxBlockSize=20000 and other hyper-parameters were as described by Foong ^39^. Each individual was assigned a score for each gene module using moduleEigengenes(). To provide biological insight into the WGCNA gene co-expression modules, we used IPA to perform functional enrichment analysis on modules with >200 genes, as the optimal set-size for IPA core analysis (QIAGEN Inc.).

## Results

### Participant characteristics

The characteristics of this study cohort and the inception birth cohort are summarised in Table 1. A lower proportion (32%) of mothers of children with autism had a university education in comparison to mothers in the random subgroup (62%). A higher proportion of children with autism were male (67%) compared to the random sub-cohort group (50%). The mode of delivery was similar between groups.

**Table 1.** Participant characteristics.

|  | Random sub-cohort group |  | Autism case group |  | Inception cohort |  |
| --- | --- | --- | --- | --- | --- | --- |
|  | N | Mean (SD) or n (%) | N | Mean (SD) or n (%) | N | Mean (SD) or n (%) |
| **Demographics and household factors (mothers)** |  |  |  |  |  |  |
| Maternal age at conception | 118 | 33.0 (4.2) | 43 | 31.0 (4.3)* | 1,064 | 31.3 (4.8) |
| Socio-Economic Indexes for Areas: | 117 |  | 42 |  | 1051 |  |
| Low (most deprived) |  | 39 (33.3%) |  | 15 (35.7%) |  | 352 (33.5%) |
| Mid |  | 35 (29.9%) |  | 14 (33.3%) |  | 350 (33.3%) |
| High (least deprived) |  | 43 (36.8%) |  | 13 (31.0%) |  | 349 (33.2%) |
| Maternal university education vs no | 118 | 72 (61.0%) | 43 | 16 (37.2%)** | 1,058 | 541 (51.1%) |
| Multiparity | 118 | 74 (62.7%) | 43 | 21 (48.8%) | 1063 | 587 (55.2%) |
| Lone parent | 118 | 4 (3.4%) | 43 | 4 (9.3%) | 1061 | 43 (4.1%) |
| **Prenatal factors (mothers)** |  |  |  |  |  |  |
| Maternal cigarette smoking (any during pregnancy) | 118 | 12 (10.2%) | 43 | 7 (16.3%) | 1051 | 168 (16.0%) |
| Maternal alcohol intake (any during pregnancy) | 116 | 59 (50.9%) | 41 | 18 (43.9%) | 980 | 515 (53.0%) |
| **Birth and postnatal factors (children)** |  |  |  |  |  |  |
| Child sex: male | 120 | 60 (50.0%) | 43 | 29 (67.4%)* | 1074 | 555 (51.7%) |
| Mode of delivery: | 120 |  | 43 |  | 1074 |  |
| Unassisted vaginal delivery |  | 53 (44.9%) |  | 15 (34.9%) |  | 525 (48.8%) |
| Instrumental vaginal delivery |  | 24 (20.3%) |  | 11 (25.6%) |  | 214 (19.9%) |
| Planned caesarean section |  | 29 (24.7%) |  | 10 (23.26) |  | 179 (16.7%) |
| Emergency caesarean section |  | 14 (11.7%) |  | 7 (16.3%) |  | 156 (14.5%) |

\* $p < 0.05$ . \*\* $p < 0.01$ . Student *t*-test or chi-squared test were used (as appropriate) to compare the participant characteristics between the random cohort group to autism case group. Inception cohort characteristics are presented for reference.

### Differences in placental mRNA transcripts in children with autism

A comprehensive profile of 16,478 substantively expressed genes was available, with the proportion of reads per sample that uniquely mapped to the genome ranging from 63.90% to 83.52%, and over 10 million annotated reads for all samples^40^. Differential expression analysis identified 1,644 DEGs in the placenta of children who developed autism compared to the random sub-cohort group (FDR < 0.05; sTable 1).

Further filtering applied by ingenuity pathway analysis (IPA) determined 1,535 DEGs as ‘analysis-ready’ for IPA analysis, including genes that are curated and annotated in the Ingenuity Knowledge Base (QIAGEN Inc; Figure 1). To evaluate the relative strength of the placental transcriptome signature for autism, we compared the placental transcriptome of BIS participants with challenge-proven IgE-mediated food allergy at 1 year of age (n=52), which is a clearly defined clinical entity with well-described differences in immune phenotype at birth^41^, to random sample of the cohort without food allergy (n=115). Strikingly, the number of DEGs associated with autism was more than 26-fold the number associated with food allergy (sFigure 1).

**Figure 1.**
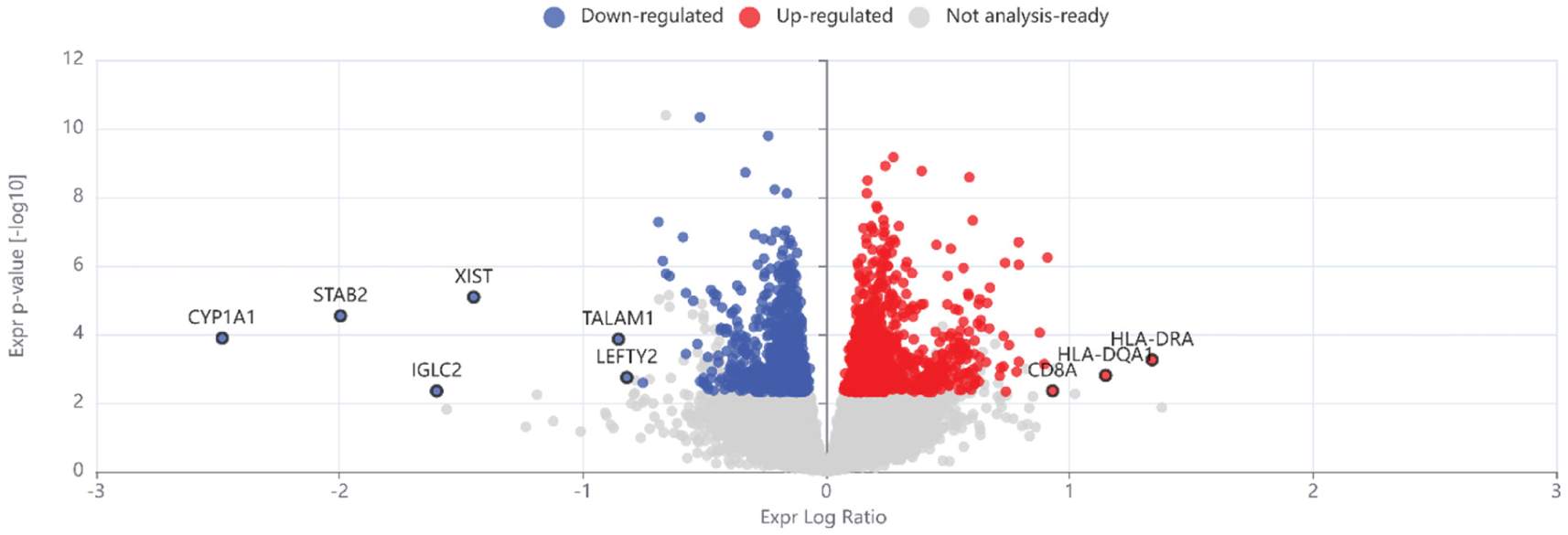
Volcano plot demonstrating placental differentially expressed genes (DEGs) in children with autism (n=43) compared with the random sub-cohort group (n=120). Negative log10 p-value (Y axis) is plotted against expression log ratio (the log2 Fold Change; X axis). Upregulated differentially expressed genes (DEGs, FDR<0.05) are shown in red and downregulated DEGs are shown in blue. Volcano plot was generated using QIAGEN’s Ingenuity Pathway Analysis (IPA).

The top upregulated DEGs were *HLA-DRA* (logFC = 1.34) and *HLA-DQA1* (logFC = 1.15), both known to regulate the major histocompatibility complex (MHC) class II and the adaptive immune response^42^ (Table 2). *CYP1A1*, encoding an enzyme of the cytochrome P450 superfamily, which is the most important xenobiotic metabolising enzyme in the placenta and regulates estrogen metabolism and lipid biosynthesis^43^, was the top downregulated DEG (logFC = −2.48). Other downregulated DEGs included *STAB2* (logFC = −1.99)*, IGLC2* (logFC = −1.60) and *XIST* (logFC = −1.45), with the latter being highly expressed in female placenta^44^ (Table 2).

**Table 2.** Top 10 differentially expressed downregulated and upregulated genes (DEGs) in children with subsequent autism diagnosis compared with the random sub-cohort group.

| Downregulated DEGs |  |  |  | Upregulated DEGs |  |  |  |
| --- | --- | --- | --- | --- | --- | --- | --- |
| Symbol | Gene Name | logFC | FDR | Symbol | Gene name | logFC | FDR |
| **CYP1A1** | cytochrome P450 family 1 subfamily A member 1 | -2.48 | 0.005 | **HLA-DRA** | major histocompatibility complex, class II, DR alpha | 1.34 | 0.013 |
| **STAB2** | stabilin 2 | -2.00 | 0.002 | **HLA-DQA1** | major histocompatibility complex, class II, DQ alpha 1 | 1.15 | 0.024 |
| **IGLC2** | immunoglobulin lambda constant 2 | -1.60 | 0.047 | **F2RL3** | F2R like thrombin or trypsin receptor 3 | 0.93 | 0.019 |
| **XIST** | X inactive specific transcript | -1.45 | 0.001 | **CD8A** | CD8 subunit alpha | 0.93 | 0.046 |
| **TALAM1** | TALAM1 transcript, MALAT1 antisense RNA | -0.85 | 0.005 | **TXNDC5** | thioredoxin domain containing 5 | 0.91 | 0.000 |
| **LEFTY2** | left-right determination factor 2 | -0.82 | 0.027 | **ARC** | activity regulated cytoskeleton associated protein | 0.90 | 0.015 |
| **PENK** | proenkephalin | -0.79 | 0.029 | **HBM** | hemoglobin subunit mu | 0.88 | 0.004 |
| **CASQ1** | calsequestrin 1 | -0.75 | 0.033 | **PRRG3** | proline rich and Gla domain 3 | 0.82 | 0.019 |
| **AGR2** | anterior gradient 2, protein disulphide isomerase family member | -0.73 | 0.029 | **TCF15** | transcription factor 15 | 0.79 | 0.000 |
| **TRNH** | tRNA-His | -0.69 | 0.000 | **SCN4B** | sodium voltage-gated channel beta subunit 4 | 0.79 | 0.014 |

A DE analysis of autism among only the male children revealed substantially concordant results to the primary analysis, and a DE analysis of the sex assigned at birth among the full analysis cohort showed no concordance with the primary analysis (sFigure 2). Therefore, sex assigned at birth did not predict the differential expression patterns for autism here.

**Figure 2.**
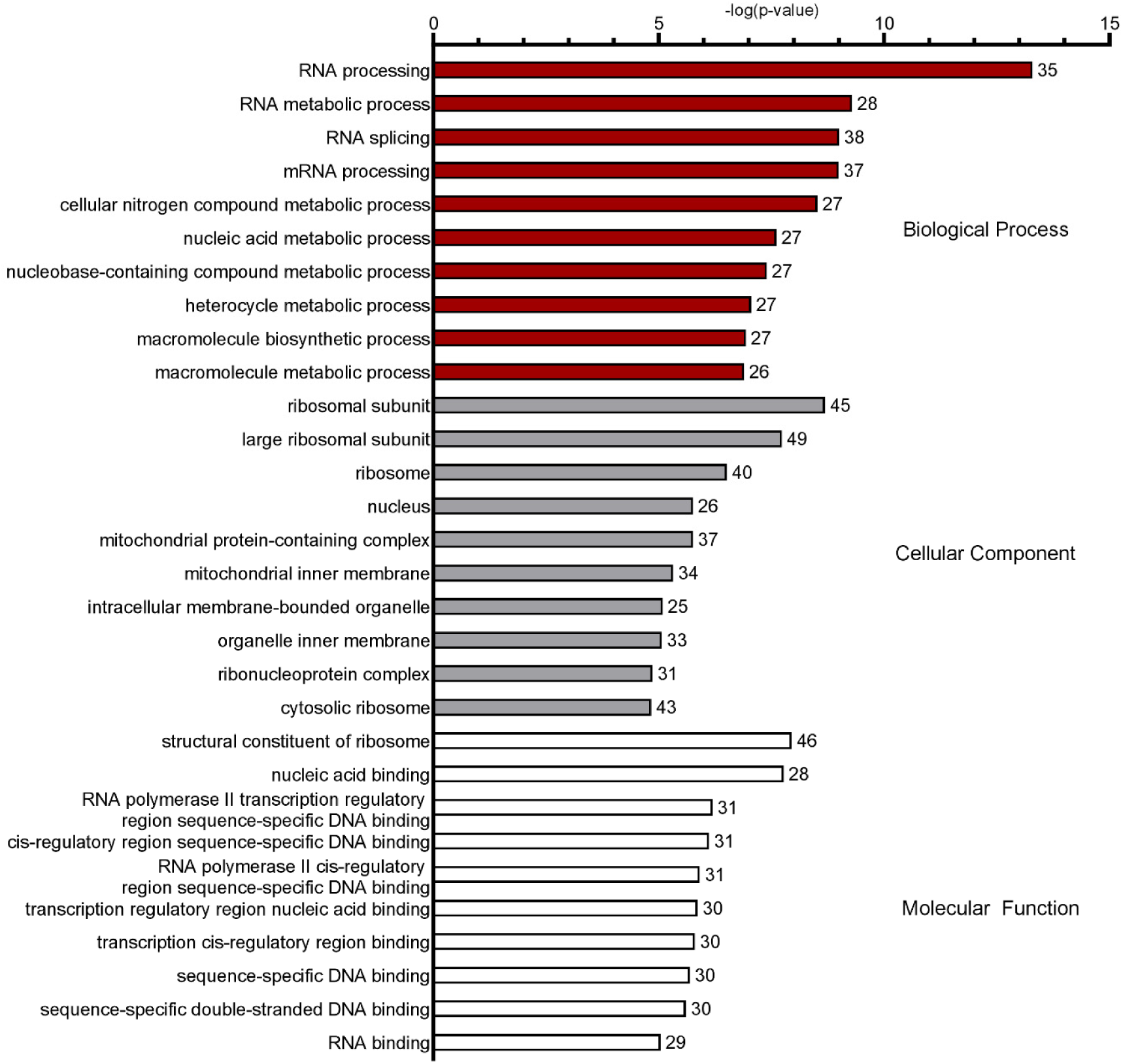
Gene Ontology (GO) enrichment analysis of the placental differentially expressed genes (DEGs) for children with subsequent autism diagnosis showing top 10 GO annotations per term. The percentage of DEGs from the total number of genes associated with the term are presented on the right-hand side of the bar.

### Gene ontology (GO) and pathway enrichment analyses

Top GO annotations associated with autism included biological processes regulating RNA processing, metabolic processes, and splicing. In the cellular component ontology, terms related to the ribosome, nucleus and mitochondria were among the most overrepresented in the DEGs. Molecular functions related to ribosomal structure, nucleic acid binding and RNA transcription were also among the top enriched terms (Figure 2; sTable 2).

Top KEGG pathways enriched for upregulated DEGs associated with autism included several pathways of neurodegenerative disease, energy metabolism and translation. Downregulated DEGs were enriched in KEGG pathways of infectious disease and transcription (Figure 3A, sTable 3A, B). Reactome enrichment analysis identified pathways of energy metabolism, as well as RNA and protein processing as being the most significantly enriched (Figure 3B, sTable 4).

**Figure 3.**
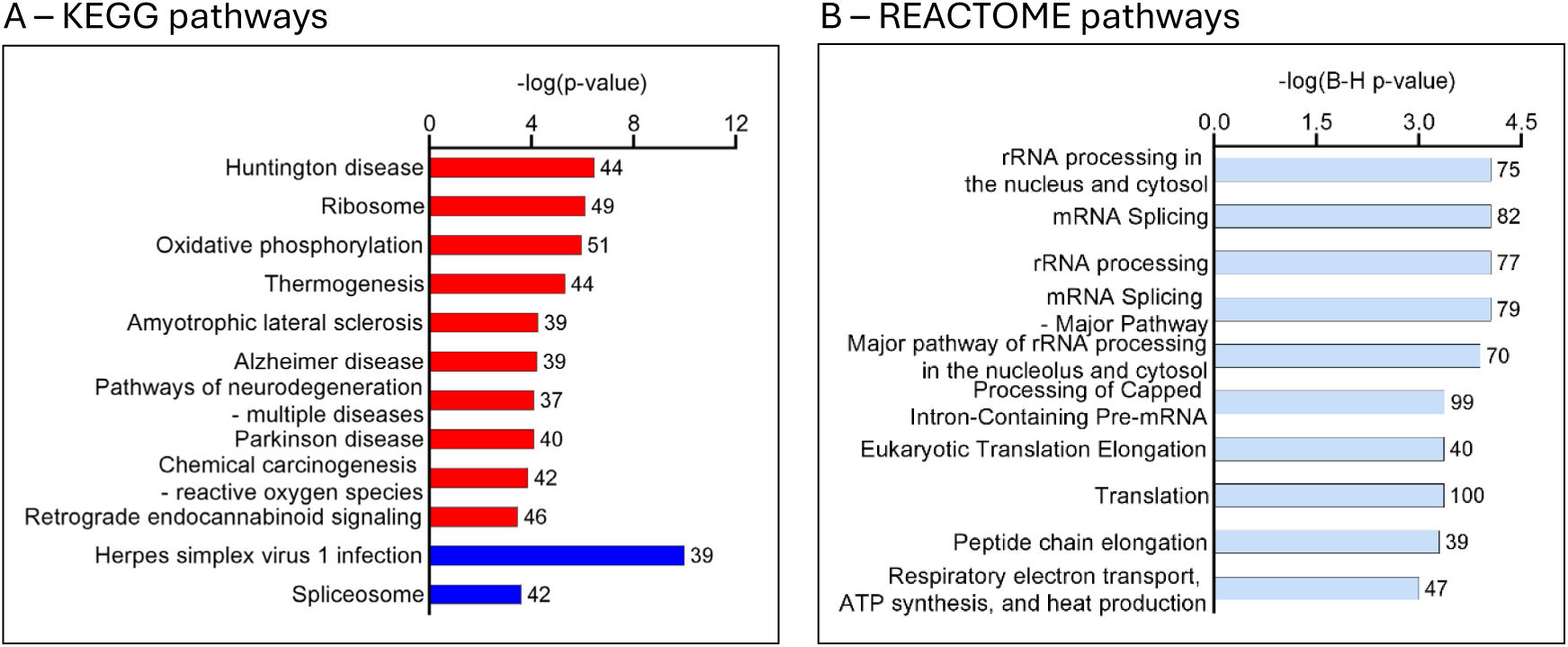
Top statistically significant enriched pathways in placental tissue from children with subsequent autism diagnosis. **(A)** Red bars indicate upregulated KEGG pathways, blue bars indicate downregulated pathways. Pathways are presented in the order of significance value (−log (p-value)) for upregulated pathways followed by downregulated pathways. Numbers on the right-hand side of the bars indicate the percentage of DEGs out of the total number of genes in the pathway. **(B)** Top 10 enriched Reactome pathways. Numbers on the right-hand side of the bars indicate the total number of genes associated with the pathway. Pathways are presented in the order of significance value (−log (BH p-value)).

### Ingenuity Pathway Analysis (IPA): Canonical pathways and upstream regulators

IPA identified 65 significantly enriched canonical pathways, representing significant activation or inhibition of a canonical pathway. Like GO annotations, KEGG and Reactome pathway analyses, the top inhibited canonical pathways related to RNA metabolism, as well as mitochondrial transfer RNA processing. Mitochondrial translation was the top activated canonical pathway, along with pathways of apoptosis and autophagy (Figure 4A). The DEGs involved in these pathways are reported in sTable 5.

**Figure 4.**
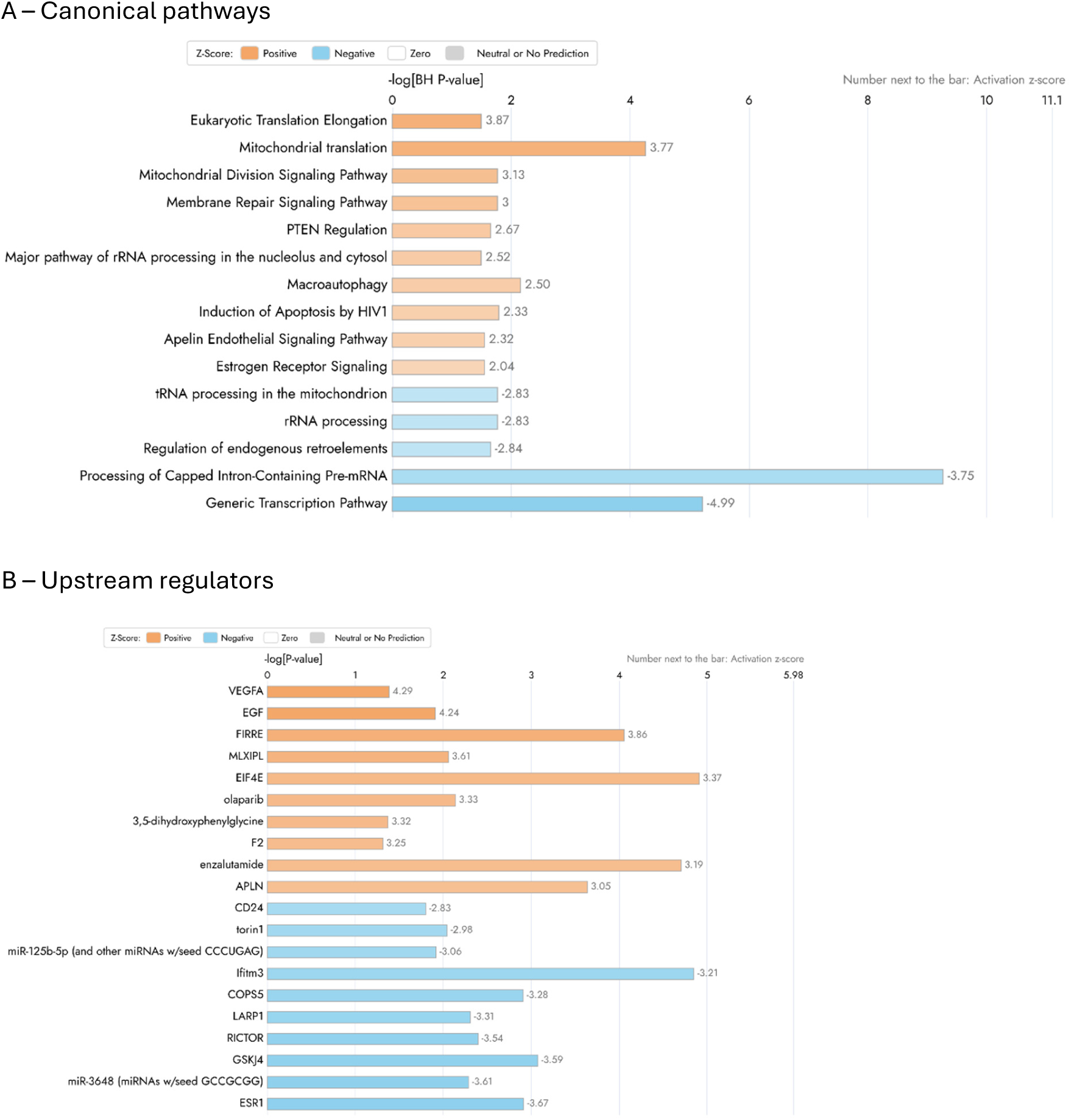
Canonical pathways and upstream regulators in placental tissue of children with subsequent autism diagnosis. Top statistically significant enriched **A)** canonical pathways, and **B)** upstream regulators in children with autism diagnosis compared with the random sub-cohort. Data are presented in the order of activation z-score (number next to the bar). For ease of depiction, only the top canonical pathways (FDR < 0.05) are presented. Orange colour range indicates activated, and blue colour range indicates inhibited pathways and regulators. Figures were generated using QIAGEN’s Ingenuity Pathway Analysis (IPA).

ESR1 (estrogen receptor 1), miR-3648 (microRNA) and GSKJ4 (a histone demethylase inhibitor) were the top upstream regulators predicted to be inhibited in the autism case-group, while VEGFA (vascular endothelial growth factor A), EGF (epidermal growth factor) and Firre (RNA element) were the top upstream regulators that were predicted to be activated (Figure 4B, sTable 6).

Causal Network Analysis further identified 192 autism-specific master regulator molecules that were predicted to influence the expression of target genes (sTable 7). The top inhibited master regulators that were predicted to be associated with autism, via the regulation of DEGs in our dataset, included SMARCA2 (transcription regulator), MNAT1 (component of the key cell cycle kinase cyclin dependent kinase 7), TFIIH (a complex that regulates RNA polymerase II transcription and other biological roles) and ESR1, while the top activated master regulators were nimesulide (a cyclooxygenase-2 inhibitor that is predicted to inhibit *CYP1A1* expression in our dataset), fluoxymesterone (anabolic androgenic steroid that has been shown to inhibit ESR1^45^, including in our dataset), Firre and THZ2 (CDK7 inhibitor known to suppress cell growth and proliferation). The top and most direct causal networks containing one intermediate regulator are presented in sFigure 3.

### Prediction of autism status

Across 100 training:test partitions we found a median sensitivity of 0.57 (2.5th–97.5th centiles: 0.29, 0.76) and median specificity of 0.92 (2.5th–97.5th centiles: 0.78, 0.96) for predicting autism (Figure 5), with median positive and negative likelihood ratios of 6.43 (2.5th–97.5th centiles: 2.86, 30.2) and 0.48 (2.5th–97.5th centiles: 0.28, 0.79), respectively. These data indicate a stronger diagnostic utility for identifying autism status rather than excluding it.

**Figure 5.**
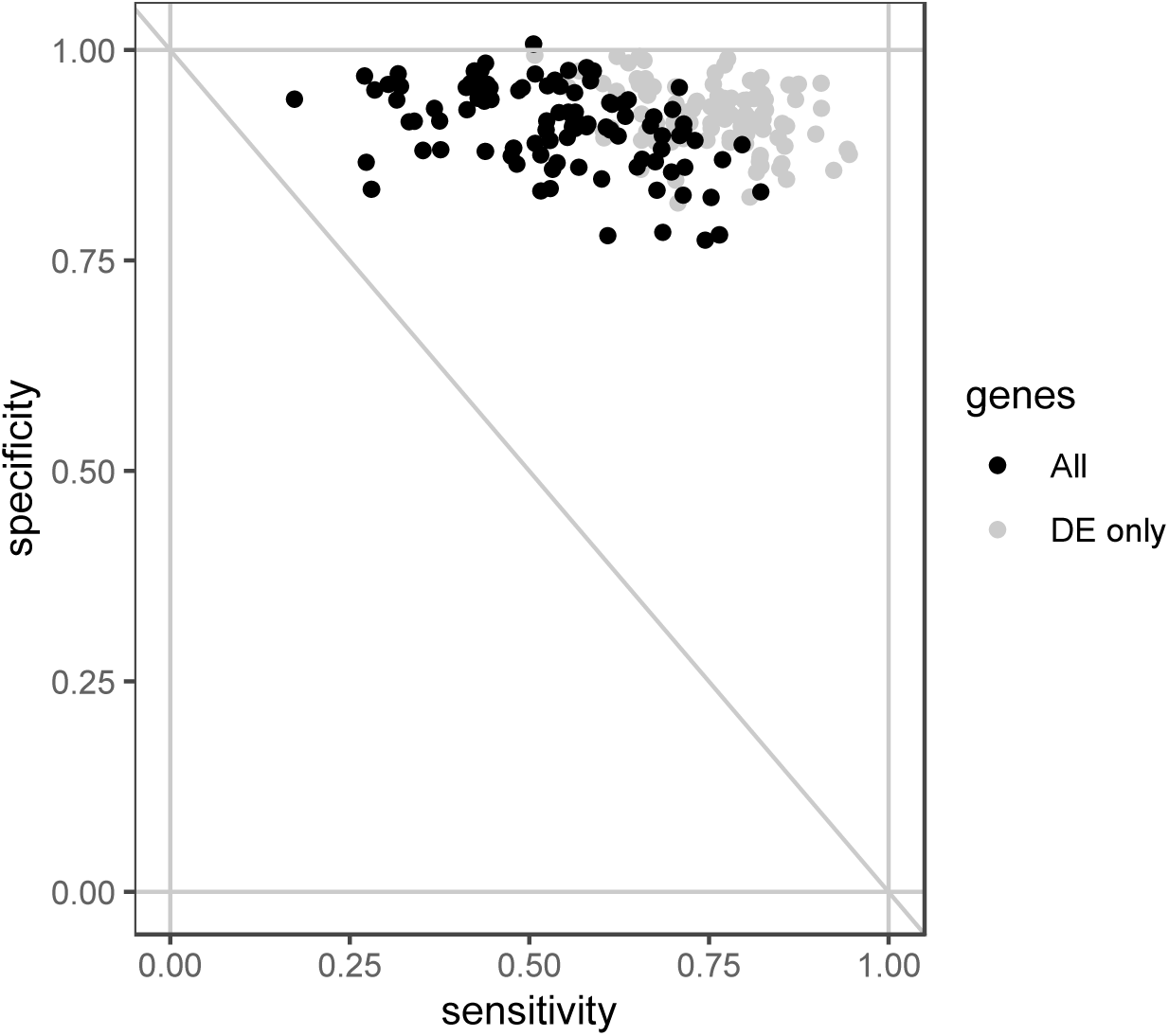
Sensitivity and specificity of sparse partial least squares discriminant analysis (sPLS-DA) for predicting autism status from placental transcriptomics and foetal sex. Black dots arise from predicting based on all 16,478 genes originally included in the differential expression (DE) analysis in autism vs. controls. Grey dots arise from prediction based only on the 1,644 DEGs (FDR<0.05). A small amount of jitter has been added to reduce overplotting and is responsible for specificity values appearing to exceed 1.

### Weighted Gene Co-expression Network Analysis (WGCNA)

WGCNA identified 8 modules of correlated genes, each identified by a colour, and a grey module comprised of genes that were not clustered in any co-expression module and was not considered further (Figure 6; sTable 7).

**Figure 6.**
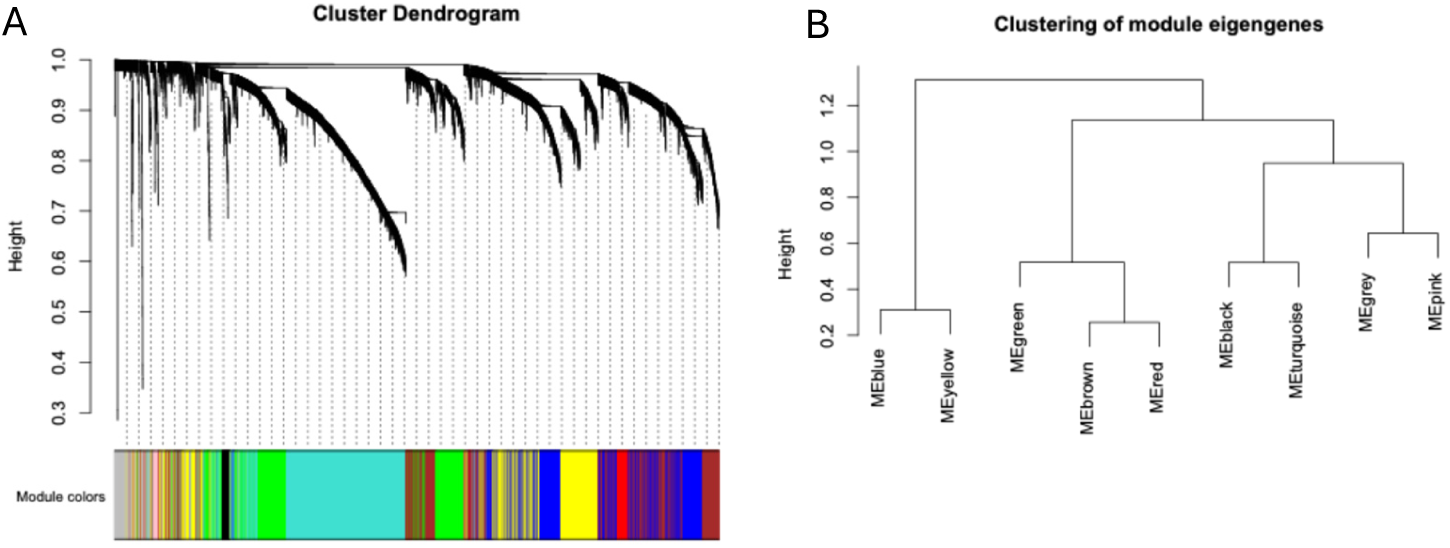
Construction of gene modules using WGCNA. *A)* The cluster dendrogram built by hierarchical clustering of the dissimilarity matrix, where dissimilarity is represented by height. Each colour represents one module and grey are unclustered genes. ***B)*** Clustering dendograms of module eigengenes for identifying meta-modules.

### Functional enrichment analysis of WGCNA modules

Given that the optimal dataset size for IPA core analysis is 200-3,000 genes, only the largest five modules with>200 genes were selected for canonical pathway analysis using the IPA. These included turquoise, green, brown, blue and yellow modules (sTable 8). The major enriched pathways in the first meta-module consisting of blue and yellow eigengenes (Figure 6B) were associated with oxidative phosphorylation and RNA transcription (sTable 8). Overall, the major themes/pathways arising from WGCNA functional enrichment were associated with mitochondrial oxidative phosphorylation and heme signalling (blue and green modules); RNA transcription and ribosome/translation (brown, yellow and green modules); ER stress and vesicular trafficking (yellow module); as well as vascular–stromal and fibrotic remodelling (turquoise module).

## Discussion

Impaired mitochondrial function is understood as a key mechanism linking exposure to specific prenatal environmental factors to autism risk^46,47^. Here, we report differential expression of genes and enrichment of gene pathways and gene co-expression modules regulating mitochondrial function in the placenta at birth of infants subsequently diagnosed with autism. *CYP1A1*, a key enzyme for metabolising both endogenous substrates, such as hormones, and xenobiotics^43^, was the top downregulated DEG in the placenta of children with a subsequent autism diagnosis. Reduced capacity of the placenta to metabolise endogenous substrates, xenobiotics, and other exogenous toxicants may be a critical determinant of the impact of endocrine-disrupting chemicals, such as phthalates and bisphenols, in the maternal circulation^48,49^. Further, using a machine learning approach, we demonstrated that placental transcriptome could predict autism with a high level of specificity in this setting, thus identifying high-risk infants for future intervention studies.

The placenta expresses metabolising enzymes that regulate foetal exposure to steroid hormones, vitamins, fatty acids, drugs, environmental chemicals and other compounds found in the maternal circulation^50^. Placental metabolism of these substrates is critical for foetal brain development^11^. CYP450 (CYP) is the most well-characterised enzymatic system involved in placental xenobiotic and drug metabolism as well as biosynthesis and degradation of endogenous substrates that are vital for foetal growth. CYP enzymes are found in the mitochondria and endoplasmic reticulum of placental trophoblast cells^51,52^. The expression of placental CYP enzymes varies during gestation, however CYP1A1 is detectable throughout pregnancy^50,52^. CYP1A1 metabolises many xenobiotics and endogenous substrates, such as estrogens, melatonin, arachidonic and eicosapentaenoic acid, which are critical for foetal development^53^. The expression of CYP1A1 steadily increases during pregnancy, and its inhibition or knockout is associated with embryonic/foetal lethality and/or developmental defects in preclinical studies^54,55^. A virtual screening model for CYP1A1 inhibitors has identified several suspected endocrine disruptors, such as bisphenol A and phthalates, acting as potent inhibitors of CYP1A1^53^. We recently demonstrated that maternal prenatal bisphenol A levels are associated with an elevated risk of autism diagnosis in boys through pathways involving inhibition of another CYP enzyme, brain aromatase (CYP19A1)^56^. In addition to inhibition of CYP enzymes, bisphenol A can cause oxidative stress, aberrant mitochondrial biogenesis and bioenergetics, decreased mitochondrial membrane potential, mitophagy, and apoptosis^57^. Phthalates are similarly toxic to mitochondria and are associated with autism risk^58^, particularly among infants with high genetic scores for oxidative stress vulnerability^59^. Thus, the interplay between ambient environmental exposures and genetic risk leading to impaired reactive oxygen species (ROS) processing may be crucial in autism pathogenesis^59^, with mitochondria being the major contributor to ROS production^60^. A placental transcriptome signature indicative of both xenobiotic susceptibility and impaired mitochondrial function among infants subsequently diagnosed with autism has, therefore, implications for mechanistic understanding, and for underpinning future research targeting the prenatal window of autism development and may also be relevant to the reported increase in autism prevalence.

One key factor in regulating CYP1A1 is the aryl hydrocarbon receptor (AHR)^61^. Exogenous environmental contaminants and dietary compounds primarily induce activation of AHR^43^. However, AHR also responds to endogenous ligands, including products of the tryptophan/kynurenine pathway^62^, and ligand-independent activation of AHR has also been documented (reviewed in ^63^). Activation of AHR leads to translocation of AHR to the nucleus, usually inducing CYP1A1 expression and its effects on metabolism and detoxification of environmental toxicants. Disruption to these usual activation processes may, therefore, also contribute to the downregulated expression and activity of CYP1A1, leading to increased bioavailability of environmental, dietary, therapeutic and other compounds, potentially resulting in adverse health effects^64^.

Placental CYP1A1 activity appears to be induced by maternal smoking and alcohol consumption^65,66^ and reduced by maternal obesity^67^. The use of selective serotonin reuptake inhibitors (SSRIs) during pregnancy has also been associated with reduced CYP1A1, as well as reduced CYP19A1 (aromatase) activity in the placenta^68^. While in the current study, placental expression of *CYP19A1* mRNA was not significantly altered in association with autism, the IPA causal network analysis predicted significant inhibition of estrogen receptor alpha (*ESR1*), as one of the top upstream regulators associated with autism, via the regulation of DEGs in our dataset. Estrogen synthesis is catalysed by CYP19A1 and placental ESR1 mediates the stimulatory effects of estrogen on *CYP19A1* expression and activity in the human placenta^69^. Overall, our data suggest that changes in placental metabolism may mediate the impact of environmental exposures on foetal neurodevelopment, with a possible involvement of placental aromatase activity in this relationship.

The top upregulated DEGs in our dataset were immuno-regulatory genes, including the HLA genes, *HLA-DRA* and *HLA-DQA1, F2RL3* and *CD8*. The HLA genes/haplotypes are instrumental for immune responses and are also implicated in autism risk^70–72^. Given the complexity and diversity of the HLA system, the mechanistic link between the HLA genes and autism risk is poorly understood^73^. Recent evidence suggests that at least some HLA variants may mediate the association between gut inflammation and autism^70^. However, future research is required to understand the links between variations in HLA genetic diversity, including in its placental expression, and subsequent autism risk. Notably, these top upregulated DEGs were not enriched in significant GO terms or gene pathways, with the top downregulated herpes simplex virus 1 infection KEGG pathway driven entirely by the differential expression of zinc-finger proteins, a superfamily of transcription factors^74^. This is likely because there were relatively few immune-related DEGs in our study^75^.

The DEGs from the placenta of infants with a subsequent autism diagnosis were enriched in pathways related to mitochondrial function, oxidative stress and ribosomal signalling in the pathway and WGCNA analyses we performed. Predicted upstream regulators also included molecules associated with RNA processing in the mitochondria, transcription and translation. Mitochondrial function tightly regulates placental transcriptional responses and placental metabolic activity^76^. Unsurprisingly, mitochondrial dysfunction in the placenta and placental mtDNA mutational load are associated with impaired foetal growth^77^. Exposure to adverse environmental conditions during pregnancy, including poor nutrition, environmental toxins, smoking, obesity and pregnancy complications associated with hypoxia, have all been associated with placental mitochondrial dysfunction and placental insufficiency^78^.

Beyond infancy, individuals with autism appear to have altered mitochondrial metabolism, with elevated blood and urine concentrations of metabolites and enzymes linked to mitochondrial function, including lactate, pyruvate, alanine and creatine kinase^79^. It is plausible that persisting mitochondrial dysfunction in individuals with autism originates from prenatal exposures to any of a multitude of toxicants, including cigarette smoke, pollution, plasticisers, bisphenols, inflammation, medications such as selective serotonin reuptake inhibitors and nutritional deficiencies^47,80,81^. We have previously shown that increased prenatal phthalate exposure is associated with autism symptoms at 2 years of age^82^. This association was partially mediated by a metabolic shift in maternal energy metabolism to non-oxidative glycolytic metabolism, indicated by increased maternal blood concentrations of lactate and pyruvate^82^. Further research is required to understand the role of placental mitochondrial function in relation to prenatal environmental exposures and autism risk. Impaired placental mitochondrial function may be a common pathway mediating the association between multiple adverse prenatal environment factors and autism development.

In addition to mitochondrial-specific pathways, the top GO biological process annotations included terms associated with RNA processing and metabolism. Terms annotated under this GO category are reflective of larger biological programs and therefore in addition to mitochondria, these processes can also take place in the cytoplasm and the nucleus^83,84^. RNA processing has an essential role in neurodevelopment, and its disruption has been previously documented in individuals with autism and in neurodegenerative conditions^84^. Notably, the top upregulated KEGG pathways in association with autism included those previously associated with neurodegenerative diseases, such as Huntington’s, Alzheimer’s and Parkinson’s diseases. While the issue of neurodegeneration in autism is debated^85^, several studies show evidence of progressive neurodegeneration in the brains of individuals with autism^86,87^, including a higher rate of parkinsonism and Parkinson’s disease diagnosis in autistic adults, compared with the general population^88–90^.

The strengths of this study include the nested case-cohort sample with prospective data within a large prebirth cohort study, assembled using an unselected sampling frame. We collected placental samples in >95% of BIS infants; the 9-year retention rate was 85% and we were able to review the relevant medical records of 99% of children with parent-reported autism or autism under assessment. Two experienced BIS clinicians validated that a paediatrician had diagnosed the children with autism against DSM-5 criteria. Further, our findings are consistent with previous evidence in BIS regarding a shift to glycolytic energy generation with elevated lactate and pyruvate during pregnancy^82^ and the mounting literature regarding xenobiotics and mitochondrial function in autism^79–81^. We were able to detect relevant biological pathways agnostically, subsequently confirming the major mitochondrial and RNA processing-related pathways as also acting within co-expression gene modules.

One limitation of this work is the use of bulk RNA-seq data, rather than single-cell RNA-seq, which enables the assessment of gene expression levels across different cell types. While the latter approach would be informative in heterogenous tissue such as the placenta, our analysis included tissue biopsies taken from different placental regions, with each sample being an aggregate measure of placental gene expression. It is also noteworthy that bulk RNA-seq generates a better overall view of the transcriptional landscape^91,92^. Hence, future autism placental studies should incorporate single-cell RNA-seq or spatial transcriptomics to understand the role of specific cell types. Future work could also consider additional environmental and biological data to explore mediation pathway. Proteomics and *ex-vivo* metabolic studies are required to confirm the functional correlates of the observed gene transcriptome signature.

Importantly, we employed sPLS-DA, a multivariate dimensionality-reduction machine learning tool^36^, to predict autism based on placental gene transcription. Our findings demonstrating a median sensitivity of 57% and median specificity of 92% are based on a case-cohort subgroup sample of the BIS cohort, including 43 children with a validated autism diagnosis and 120 children randomly selected from the remainder of the cohort. A single sPLS-DA prediction model can be derived from these 163 samples for deployment in other cohorts, either prospectively or in birth cohorts with stored placental samples and children with subsequent autism diagnosis. The high specificity highlights the importance of prenatal factors for autism onset. The moderate sensitivity may be because a subgroup of children required additional postnatal factors to precipitate autism. Prediction modelling based on the placental transcriptome may enable the identification of high-risk infants for future intervention studies to improve autism outcomes.

In conclusion, our findings show that placental tissue collected at birth from infants with a subsequent diagnosis of autism has distinct changes in genes and gene subsets involved in xenobiotic metabolism, mitochondrial function and RNA processing. These findings add to the growing body of literature implicating impaired mitochondrial function as a possible key mechanism in autism development. Future research should investigate the drivers of such mitochondrial function, prioritising organic pollutants such as bisphenol A and phthalates^93^, as well as the functional implications of these placental transcriptional changes and their role in the association between prenatal environmental exposures, epigenetic mechanisms and autism development and diagnosis.

## Supporting information

sTable

sFigure

## Data Availability

Barwon Infant Study (BIS) data requests are considered on scientific and ethical grounds by the BIS Steering Committee. If approved, data are provided under collaborative research agreements.

## Acknowledgements

We thank the study participants, as well as the entire BIS team, including interviewers, nurses, computer and laboratory technicians, clerical workers, research scientists, volunteers, managers, and receptionists. We also thank the obstetric and midwifery teams at Barwon Health and St John of God Hospital Geelong for their assistance in recruitment and collection of biological specimens.

## Funding

The establishment work and infrastructure for the BIS was provided by the Murdoch Children’s Research Institute, Deakin University, and Barwon Health, supported by the Victorian Government’s Operational Infrastructure Program. Funding was also provided by the National Health and Medical Research Council of Australia (NHMRC), the Jack Brockhoff Foundation, the Shane O’Brien Memorial Asthma Foundation, the Our Women’s Our Children’s Fund Raising Committee Barwon Health, The Shepherd Foundation, the Rotary Club of Geelong, the Ilhan Food Allergy Foundation, GMHBA Limited, Vanguard Investments Australia Ltd, and the Percy Baxter Charitable Trust, Perpetual Trustees, and Minderoo Foundation. In-kind support was provided by the Cotton on Foundation and CreativeForce. This work was also supported by NHMRC - EU Joint Programme – Neurodegenerative Disease Research (JPND) and the RANZCP foundation Pat, Toni and Peter Kinsman Research Scholarship to LS. DB is supported by a NHMRC Leadership 1 Investigator Grant (GTN1175744). MB is supported by a NHMRC Leadership 3 Investigator grant (GNT2017131).

## Author contributions

L.S., A.L.P., M.O’H., R.S., P.D. and P.V. conceived of the study and designed the protocol. L.S. and R.S. conducted and supervised the laboratory work. L.S., M.O’H. and S.T., conducted the statistical analyses. L.S. and P.V. wrote the original draft of the paper. A.L.P., M.O’H., R.S., C.S., P.D., D.B., P.D.S., F.C., S.T., K.D., C.J.L., T.M., K.V., S.L.M. and M.B. critically revised successive drafts of the paper and approved its final version.

## Competing interests

A.L.P. and P.V. have a financial interest in Meizon Innovation Holdings Pty Ltd.

## Notes

### Author Declarations

The study was approved by the Barwon Health Human Research Ethics Committee (HREC 10/24), and families provided written informed consent.

