## Supplementary material for "A placental transcriptional signature for autism": sFigure

**Supplementary Figure 1. Volcano plot demonstrating placental differentially expressed genes (DEGs) identified in children with challenge-proven IgE-mediated food allergy at 1 year of age (n=52) compared with a random sample of the cohort without food allergy (n=115).** Among 16,241 genes with expression values, differential expression analysis identified a statistically significant difference in the expression of 57 genes (FDR < 0.05).

Full details on food allergy diagnosis protocols and methods in the BIS cohort have been described previously (Zhang, Y., et al., *Cord blood monocyte-derived inflammatory cytokines suppress IL-2 and induce nonclassic "T(H)2-type" immunity associated with development of food allergy.* Sci Transl Med, 2016. **8**(321): p. 321ra8.).

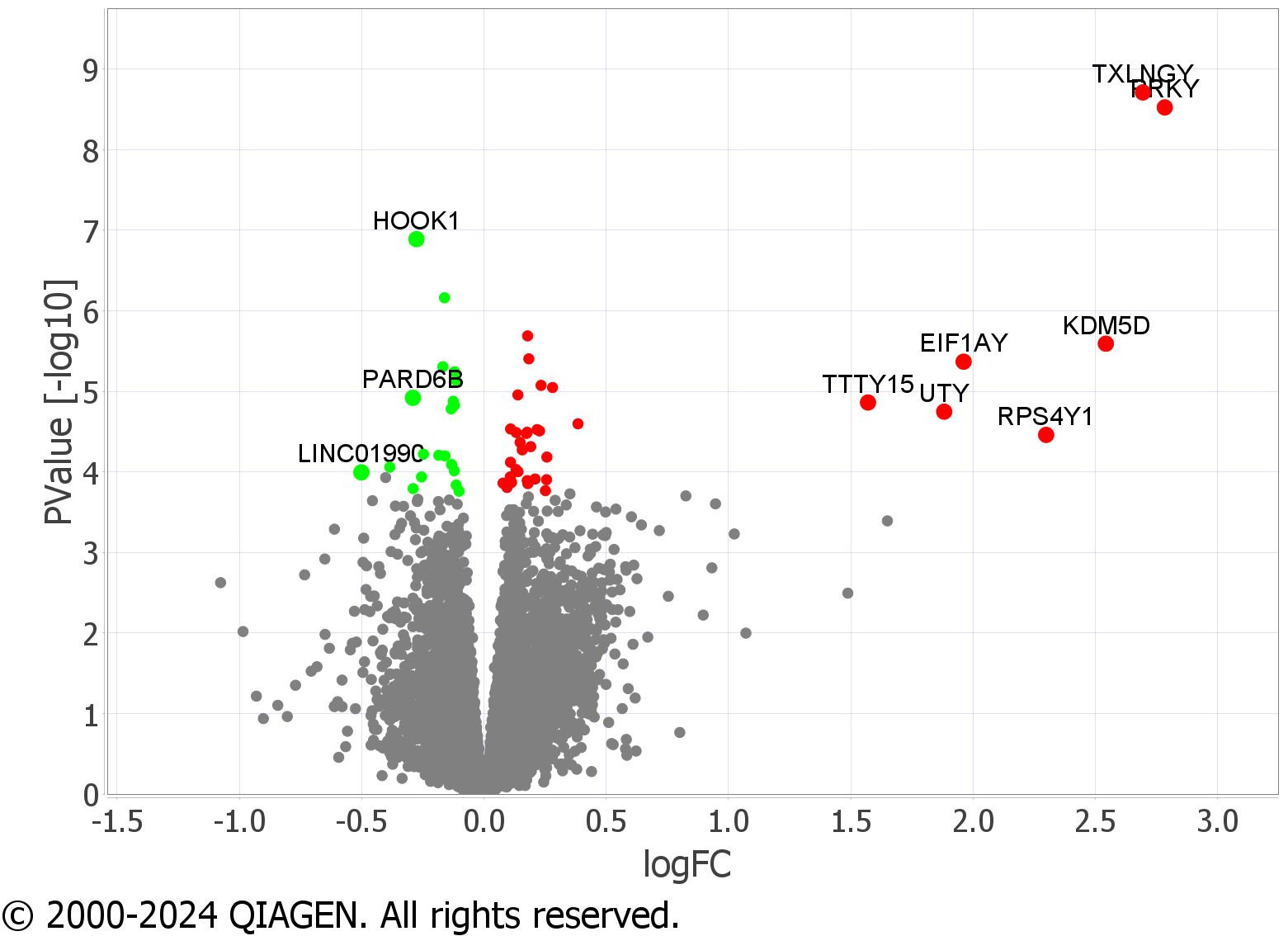

**Supplementary Figure 2.** **Plots comparing primary and sensitivity differential expression (DE) analyses**. DE analysis of autism spectrum disorder (ASD) adjusted for infant sex from the main text, with **(A,C)** DE analysis of ASD among male infants only; and with **(B,D)** DE analysis of infant sex among all subjects. Venn diagrams **(A,B)** show the numbers of genes found to be differentially expressed at a false discovery rate of 0.05 in the two analyses. Hypergeometric tests suggested by these diagrams give p-values of <0.001 and 0.9998 respectively for the intersection to contain as many genes or more under a null hypothesis that the DE genes are selected uniformly at random from all those in the original analysis. Plots of the ranks of p-values of DE genes **(C,D)**; black dots represent genes DE in both analyses, the grey are genes DE in only one analysis) show a concordance between the DE genes in the sex-adjusted ASD analysis and in the sex-specific analysis **(C)** but not between the sex-adjusted analysis and the analysis of infant sex **(D)**.

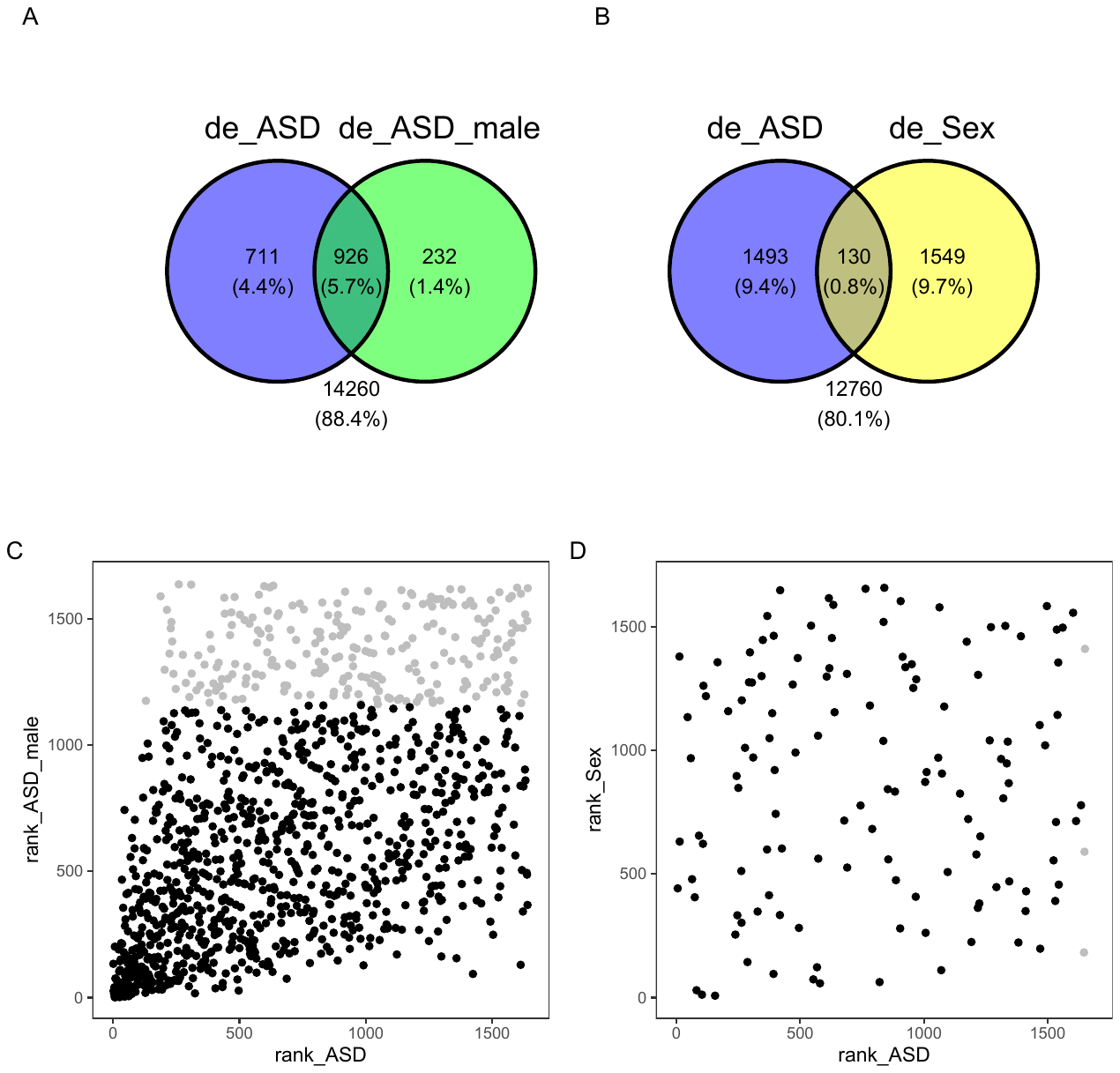

**Supplementary Figure 3.** **Visual representation of regulatory networks that represent possible causes (upstream regulators) and underlying mechanisms for observed molecular changes in the dataset (differentially expressed genes; DEGs), and their possible association with diseases and functions of interest. A)** Visualises the relationship between *ESR1* as the predicted inhibited upstream regulator that is at the top of the causal network, DEGs and ‘autism-like behavioral deficit’, a pre-defined term based on the QIAGEN knowledgebase. **B)** Visualises the relationship between *FIRRE* as the predicted activated upstream regulator that is at the top of the causal network, DEGs and ‘autism-like behavioral deficit’. Prediction Legend provides a key to the main features of the networks, including molecule shapes and colours as well as relationship labels and types. Figures created using the Ingenuity Pathway Analysis (QIAGEN Inc).

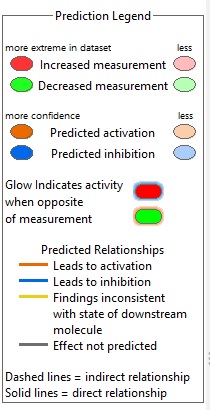

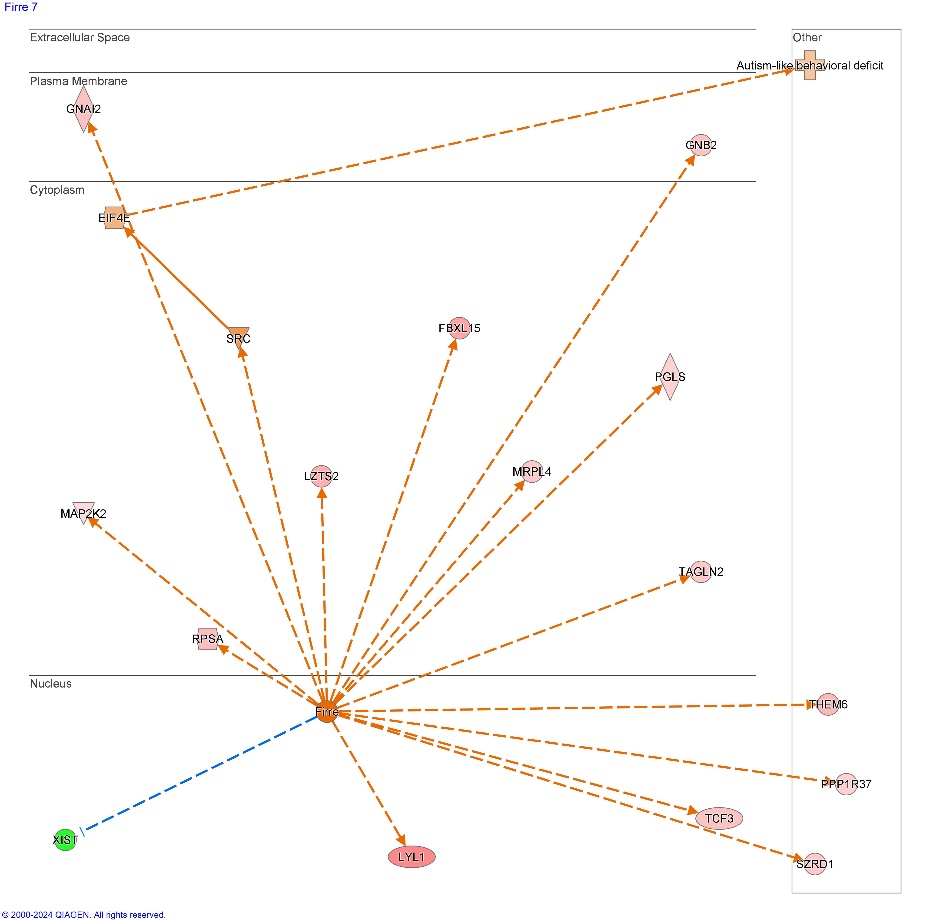

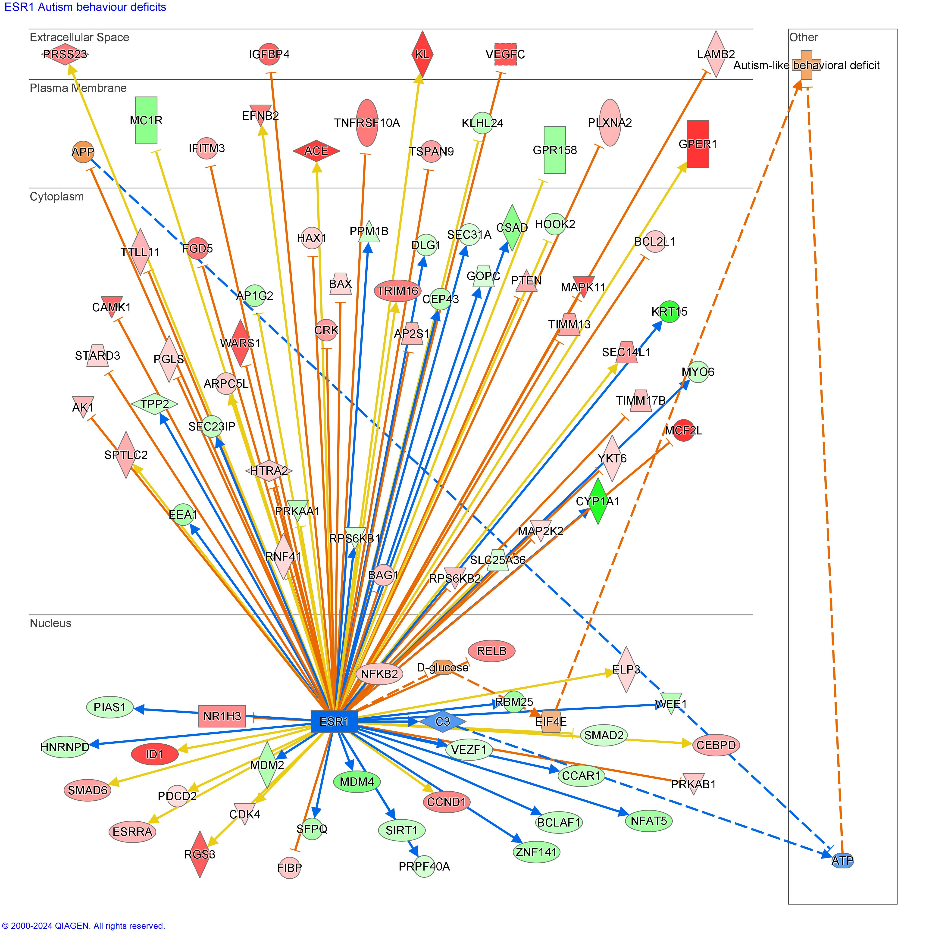
A. B.
